# Text-based Integration of Mutational Profiles for Classification and Biomarker Identification of Intraductal Papillary Mucinous Neoplasms of the Pancreas

**DOI:** 10.1101/2023.06.08.23291156

**Authors:** Nam Nguyen, Jamie K. Teer, Margaret A. Park, Francisca Beato, Patricia McDonald, Jason B. Fleming, Jennifer B. Permuth, Kwang-Cheng Chen, Aleksandra Karolak

**Affiliations:** Department of Machine Learning, H. Lee Moffitt Cancer Center and Research Institute, Tampa, FL; Department of Electrical Engineering, University of South Florida, Tampa, FL; Department of Biostatistics and Bioinformatics, H. Lee Moffitt Cancer Center and Research Institute, Tampa, FL; Department of Gastrointestinal Oncology, H. Lee Moffitt Cancer Center and Research Institute, Tampa, FL; Department of Cancer Physiology, H. Lee Moffitt Cancer Center and Research Institute, Tampa, FL; Department of Cancer Epidemiology, H. Lee Moffitt Cancer Center and Research Institute, Tampa, FL

**Author notes:** **Corresponding authors** Aleksandra Karolak Nam Nguyen. **Contributions** NN and AK conceptualized the methodology, and NN performed numerical analyses. NN and AK prepared the manuscript. NN, JKT, MAP, FB, PMD, JBF, JBP, KCC, and AK discussed the results, reviewed, and edited the manuscript.

## Abstract

**Motivation:** Intraductal Papillary Mucinous Neoplasms (IPMNs) are a common cystic precursor for pancreatic ductal adenocarcinoma (PDAC). Detecting these pre-malignant lesions poses a challenge for diagnostic tools due to their relatively low occurrence rate. However, a better understanding of the lesions’ composition could enable effective decision-making, risk assessment, treatment selection, and, most importantly, prevention.

**Methods:** In this work, we introduce a new framework for integrating information from mutational profiles using transformer-based models for stratification and biomarker identification in IPMNs vs. PDAC. We show that the numerical descriptor vectors can be used to construct highly predictive Artificial Neural Networks for disease classification. The derived mutational representations can be supported by other data types (here, mRNA) and further improve the accuracy of the classifiers. Besides the AI-driven methodology for biomarker discovery in cancer research, we also propose methods to maximize AI’s utility by recycling its knowledge to facilitate our limited understanding of the disease. We propose Natural Adversary Analysis – an AI-driven inference to detect IPMNs with a high probability of progression to malignancy.

**Results:** The proposed model supports 12 clinically relevant genetic biomarkers with high mutation rates (such as *KRAS, GNAS, ARID1A*, and *CDKN2A*) and suggests biomarkers not yet recognized (such as *RADIL, TTN*, and *ZNF287*). We broaden the study’s scope by investigating rarely mutated genes and reveal 14 biomarkers with potential clinical importance. Several genes with low mutation rates, including *TMPRSS1, CDH22, CCND2, CYFIP2, CBLL1*, and *OPCML*, are also addressed as potential biomarkers by our framework. Finally, the predictive robustness of the identified biomarker set is validated externally on the patient data from the Moffitt Cancer Center study, including six pairs of matched tumor and normal IPMN samples. We show that the presented mutational profile (MP-derived) gene panel has equivalent predictive power to clinically driven panels.

**Conclusions:** Here, we show the proof-of-concept that AI can serve the clinic and discover biomarkers beyond clinically known regimes. In line with that, we propose a translational AI-based approach for 1) disease stratification (IPMNs vs. PDAC), 2) biomarker identification, and 3) transferring the model knowledge to predict cysts’ risk of progression.

## 1) Introduction

Pancreatic ductal adenocarcinoma (PDAC) is the third leading cause of cancer-related deaths in the United States, expected to become the second by 2030 (1). The five-year survival rate is only about 11% (2). Reliable diagnostics supporting clinical decision-making for patients with increased risk for PDAC can be based on the classification of cysts. The most common types of pancreatic cysts include i) pseudocysts, which are the most common type and typically develop as a complication of acute or chronic pancreatitis; ii) serous cystadenomas - benign cysts, composed of thin-walled, multiple small compartments; iii) mucinous cystic neoplasms - typically seen in middle-aged women, characterized by thick, mucin-filled tissue and considered premalignant, with a potential risk of developing into pancreatic cancer; iv) solid pseudopapillary neoplasms - rare pancreatic cystic tumors that predominantly affect young women, typically well-defined, solid masses with cystic components; v) cystic pancreatic neuroendocrine tumors, which are another example of rare pancreatic tumors that arise from neuroendocrine cells within the pancreas; finally, vi) Intraductal Papillary Mucinous Neoplasms (IPMNs), which are characterized by the presence of papillary growths within the pancreatic ducts.

IPMNs are recognized precursors of PDAC (3). Each IPMN lesion carries 10-25% risk of progression to cancer within ten years of detection. Despite being precursors of PDAC, there is a lack of validated intervention targets for precision prevention for IPMNs; the clinical ability to stratify the risk of cancer progression of individual IPMN tumors is poor, and essentially no effective non-surgical interventions exist. Therefore, the improved detection and identification of biomarkers and pathways for malignant potential of IPMNs and their progression to PDAC to support clinical decision-making and design personalized treatments are urgently needed.

A meta-analysis of the three intrinsic genetic mutations in *KRAS, GNAS*, and *RNF43* showed the presence of these biomarkers in IPMNs as stratified by histologic grade (4). To date, *KRAS* and *GNAS* are two common clinically recognized biomarkers with the support of non-neural-based methods, such as conventional statistical tests (5). *KRAS* mutations were found in 58%, 70%, and 63% of high-grade, intermediate-grade, and low-grade IPMN samples. However, the statistical evidence did not differentiate these histologic groups by *GNAS* mutation. Moreover, due to limited data, the associations between *RNF43* and the clinicopathology of IPMNs are inconclusive in the meta-analysis. A study on the progression pathway from IPMNs to PDAC using molecular features proposes *KRAS, TP53, SMAD4*, and *CDKN2A* as *PDAC* intrinsic biomarkers, while *GNAS* mutations are often seen in IPMNs (6). A previous study also showed that the intra-tumor expression of *CES2* can sustain *HNF4* to promote PDAC progression (7). In addition, we have previously investigated mutations of potential biomarkers for PDAC, including *PTEN, PIK3CA, CTNNB1, MEN1, IDH1*, and *VHL*, among 97 preoperative pancreatic cysts (8). We found that the mutations of *GNAS* or/and *BRAF* occur in all IPMNs with 100% specificity.

Despite adding significant insights into cancer diagnosis and treatment, clinically driven biomarkers remain challenging and costly to be identified due to several reasons beyond the strict clinical protocols (9, 10, 11). First, *in vivo*, or *in vitro* validations for identified biomarkers are commonly expensive and time-consuming. Second, tumor heterogeneity adds to the complexity of this problem. Finally, validating new biomarkers meet a bottleneck of data availability, in which clinical-driven approaches are typically limited to small or intermediate cohorts. Artificial Intelligence (AI) can serve as a complementary tool to mitigate these existing problems; *In silico* strategies for biomarker discovery are cost-efficient and AI-enabled tools can identify biomarkers from larger multimodal data. Specifically, AI models can generalize their knowledge from a finite number of observations or by training on larger databases. The increasing number of publicly available cancer genomics datasets further generalize AI models. Here, transfer learning in Machine Learning (ML) and Deep Learning (DL) play key roles since knowledge from the training cohort can be hard coded into the learned model weights and transferred to a targeted cohort. A well-trained DL model allows for extracting the brief and representative numerical descriptors for given inputs that can be later used to infer some of the input’s characteristics, such as disease type or underlying pathways. This application targets clinical studies with small sample sizes by transferring useful knowledge from larger cohorts, thus improving the robustness of the biomarkers identified by ML approaches on limited data (6, 12). Regarding related works, using neural-intelligent-based methods, machine intelligence can identify data-driven biomarkers for specific cancer phenotypes (13) or biological pathways (14, 15) using neural-intelligent-based approaches.

Despite biological and clinical significance, studies addressing biomarker identification and data integration using mutational profiles are limited and technically challenging for ML. From our perspective, cancer is a causality process since only small alterations at DNA base pairs can create sequential changes in mRNA and RNA to protein functionality. Thus, the interaction terms between two inputs ***X***_Nucleotide_ (singlet of based elementary) and ***X***_Codon_ (triplet defining protein functions) need to be included for more accurate models (**Figure 1(A)**). The reason for it is that the same point-mutation C → T in a patient can lead to different mutations at the codon (amino acid) level (e.g., aCT → aTT and Cag → Tag buT Thr → Ile and Gln → Ter). Analyzing high-order interaction terms is computationally challenging for DL models due to the high dimensionality of the feature sets. Several methods have been proposed to deconvolute context-dependent mutational signatures with ML models (16, 17, 18, 19, 20, 21). The purely statistical technique relying on the matrix decomposition, the non-negative matrix factorization (NMF) method (18, 19, 20, 21, 22, 23), is commonly used to extract numeric features from the text-based mutational profiles with mutations centered at one nucleotide. This method, however, disregards the relative importance of the nucleotide mutations to alterations on the codon level **Figure 1(A)**). Although there are many publicly available datasets (24, 25, 26, 27), the existing NMF-based approaches do not address the hierarchical relationships between codon mutations and protein alterations.

**Figure 1.**
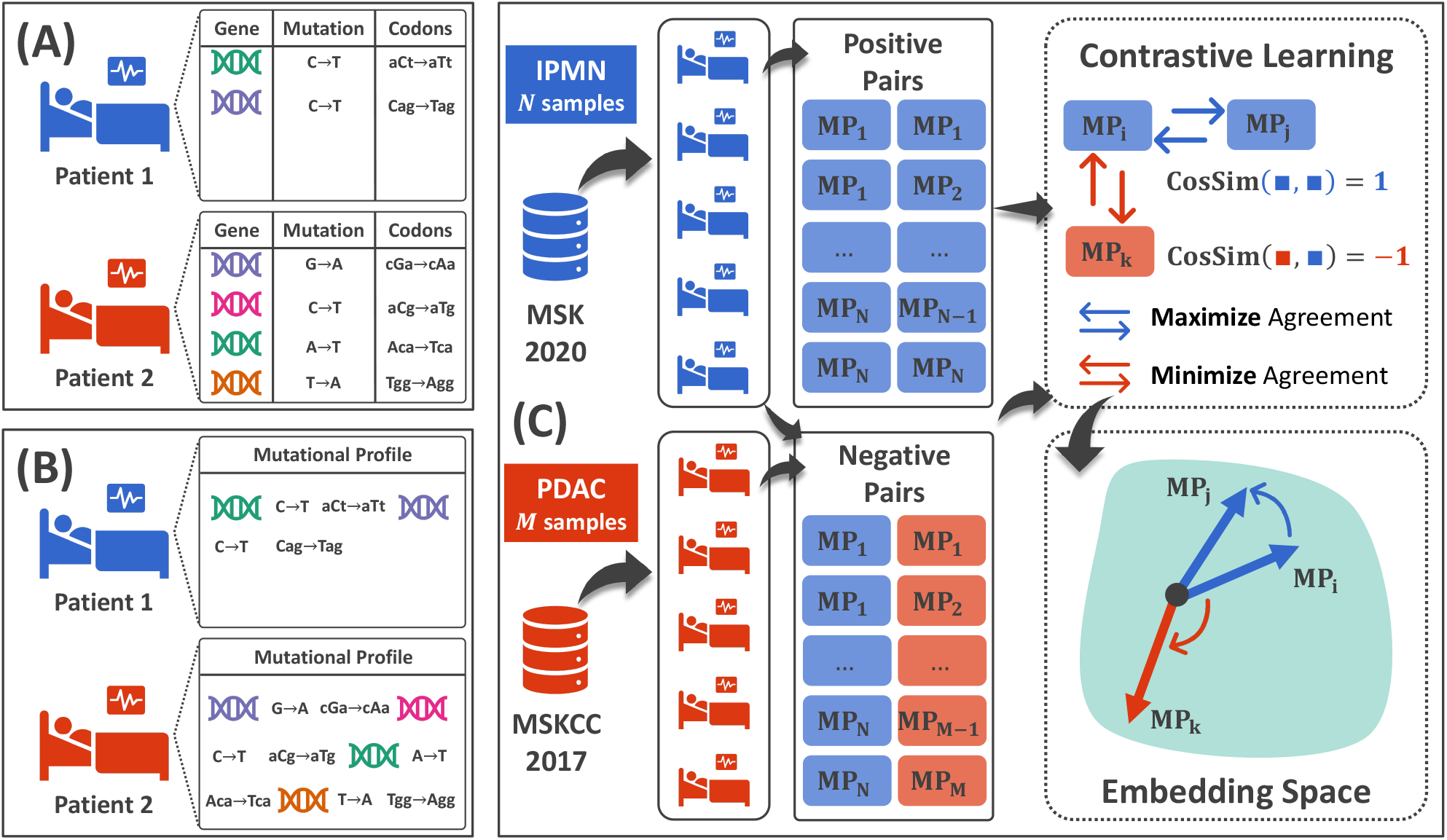
Framework of Learning Mutational Profiles Embeddings (Mutational Profile) via Contrastive Learning (CL). **(A)** Mutational information is naturally hard to be integrated into ML approaches. First, same point mutations often lead to different changes in amino acids and then protein functions. Second, there is heterogeneity in mutational profiles across patients, resulting in features of different lengths. **(B)** Our proposed processing for mutational information, in which we consider mutational profile a “language” of the human genome that could reveal the vital status of quantified patients. **(C)** CL for mutational profile embeddings. We create a large corpus of MPs based on MSK-2020 and MSKCC-2017 databases. MPs from the same class form a positive pair, while MPs from different categories form a negative pair. We consider IPMN an anchor class with *N* samples and PDAC a reference class with *M* samples. As a result, the exhaustive pairing approach derives *N* × *N* positive pairs and *N* × *M* negative pairs. CL encourages the agreement among positive pairs by maximizing their similarity (*Cosine similarity* = 1) while discouraging such an agreement among negative pairs (*Cosine similarity* = −1). As a result, mutational profile representations (or vectors) from the same disease are distinguishable from that derived from other diseases, considered representative features for ML approaches.

In this work, we introduce a new framework for integrating information embedded within the patients’ mutational profiles using transformer-based DL models. The DL neural architectures are a class of large language models that can be trained on e.g., text-based inputs to perform Natural Language Processing (NLP) tasks. Despite tremendous successes in non-medical tasks such as language translation, content generation, and text analysis, this model class has not been applied to study mutational profiles to extend our knowledge.

We develop an AI-enabled diagnostic tool for malignant phenotype stratification and biomarker identification of IPMN vs. PDAC. The central hypothesis of our work is that the mutational profile represents the natural language of the human genome that can be used to derive biomarkers for cancer phenotypes. By integrating NLP, self-attention models, and Contrastive Learning (CL), we propose to capture the intrinsic numerical descriptors from the text-based representation of mutational profiles. Thanks to the self-attention mechanism in generic transformer models, the embedded representations from the mutational profile enclose the contextual relationship between genes, nucleotides, and codon mutations. Another advantage of our framework is that the mutational profile encoders can extract features with homogenous lengths from heterogeneous text-based inputs. Thus, the retrieved numerical descriptor vectors from mutational data can be further integrated with, e.g., gene expression levels or biophysical features of proteins.

Importantly, our approach goes beyond the common somatic mutational profile analyses by utilization of the codon mutations that can further help understand the biological and clinical significance of mutations. From our perspective, codon alteration analysis could derive novel and promising biomarkers for understanding cancer development, as it provides a more comprehensive examination of the molecular changes mappable to the protein level. I.e., cancer is initiated by mutations in DNA that might lead to alterations in the structure and function of proteins. While a single point mutation (or single base substitution) analysis can address the fundamental mode of the human genome, it is often inadequate in explaining the complex changes that might occur on the cellular level. The same mutation can result in different codon configurations, amino acid changes, and protein alterations. In contrast, codon alteration analysis provides a more contextualized observation of the modifications, enabling us to gain better insights into disease development. Finally, we propose a method to recycle AI models to better understand the progression of disease (pre-malignancy to malignancy) by scoring a patient’s malignant status (progression probability) with natural adversary concepts.

We propose the Text-based Integration of Mutational Profiles for Classification and Biomarker Identification model (TI-MutaNET) we have developed here that integrates information from mutational profiles via text-based representations. To our knowledge, TI-MutaNET is the first DL diagnosis system that quantifies mutational profiles by integrating multimodality from the point mutations in DNA through codon to gene alterations. The following sections include:

- The framework of TI-MutaNET, the paradigm of stratification between IPMNs vs. PDAC, and the proposed biomarkers’ discovery method (**Section 2**).
- The results and main findings of the study (**Section 3**).
- The conclusions of the study (**Section 4**).

We discuss in Section 3 how the proposed neural pipeline can support four clinically relevant questions: 1) DL-based encoder can enable accurate ML models for classification between PDAC and IPMN samples (**Section 3.1**); 2) the proposed biomarker identification protocol enables actionable biomarkers based on point mutations, codon alteration, and gene change altogether (**Section 3.2**); 3) we construct a predictive-genetic panel for disease stratification, validated on the external dataset (**Section 3.3**). 4) the natural adversary analysis allows us to recycle optimized models to recognize IPMN samples with a high chance of progressing (**Section 3.4**).

## 2) Methods

### 2.1) Framework Overview and Datasets

#### a) Framework

To develop a model for analysis of the mutational language of the human genome, we create sentences of mutational profiles by periodically ordering gene-, nucleotide- and codon-level mutations (**Figure 1(B)**). Such data pre-processing allows us to implicitly impose the positional encoding into the sequences generated by mutational features in **Figure 1(A)**. Transformer-based models can efficiently analyze the relative importance among words (tokens) within the input sequences or mutational information **Figure 1(B)**. Attention learning allows every token to diagnose every other token’s feature, i.e., every mutational information in the input mutational profile sequence is analyzed on additional levels. **Figure 1(B)** illustrates the complexity of the mutational profile sequences, including mutated genes, nucleotide-level alterations, and amino acid changes, which can be related to protein functions. We analysed the text representations of mutational profiles using four variant neural architectures of the sentence-transformer models (28), including ALBERT (29), BERT (30), MiniLM-L6 (31) and MiniLM-L12 (31). We select the encoders with various complexity levels (ALBERT being the smallest with 11 million parameters and BERT being the largest with 110 M parameters) to quantify their effects on the final performance.

The framework of TI-MutaNET includes two steps: (1) pre-training of the mutational profile encoder (**Figure 2(A)**) and (2) leveraging encoders for final DL pipelines that can perform clinically practical tasks such as disease stratification or biomarker identification (**Figure 2(B)**). In the first step, we train the mutational profile encoder to extract representative and useful features from the text-represented mutational profile sequence. We use CL to train the encoders – this way, we simultaneously encourage the similarity (agreement) among positive pairs (same disease type) and discourage the similarity of negative paired samples (different disease types, **Figure 1(C)**). Utilizing the cosine similarity, mutational profiles from the same disease show maximum agreement (cosine similarity of 1). In contrast, the negative paired mutational profiles from different diseases will have minimized agreement (cosine similarity of −1, **Figure 1 (C)**). Thus, we encourage the mutational profile encoders to extract useful and representative features that can be used to distinguish disease conditions. We sketch the proposed CL for mutational profile encoders to extract numerical vectors on embedding vector space in **Figure 1(C)**. Importantly, starting from heterogeneous-length mutational profiles, our framework produces equal-length features controlled by the output dimensionality of the dense layer in the mutational profile encoder.

**Figure 2.**
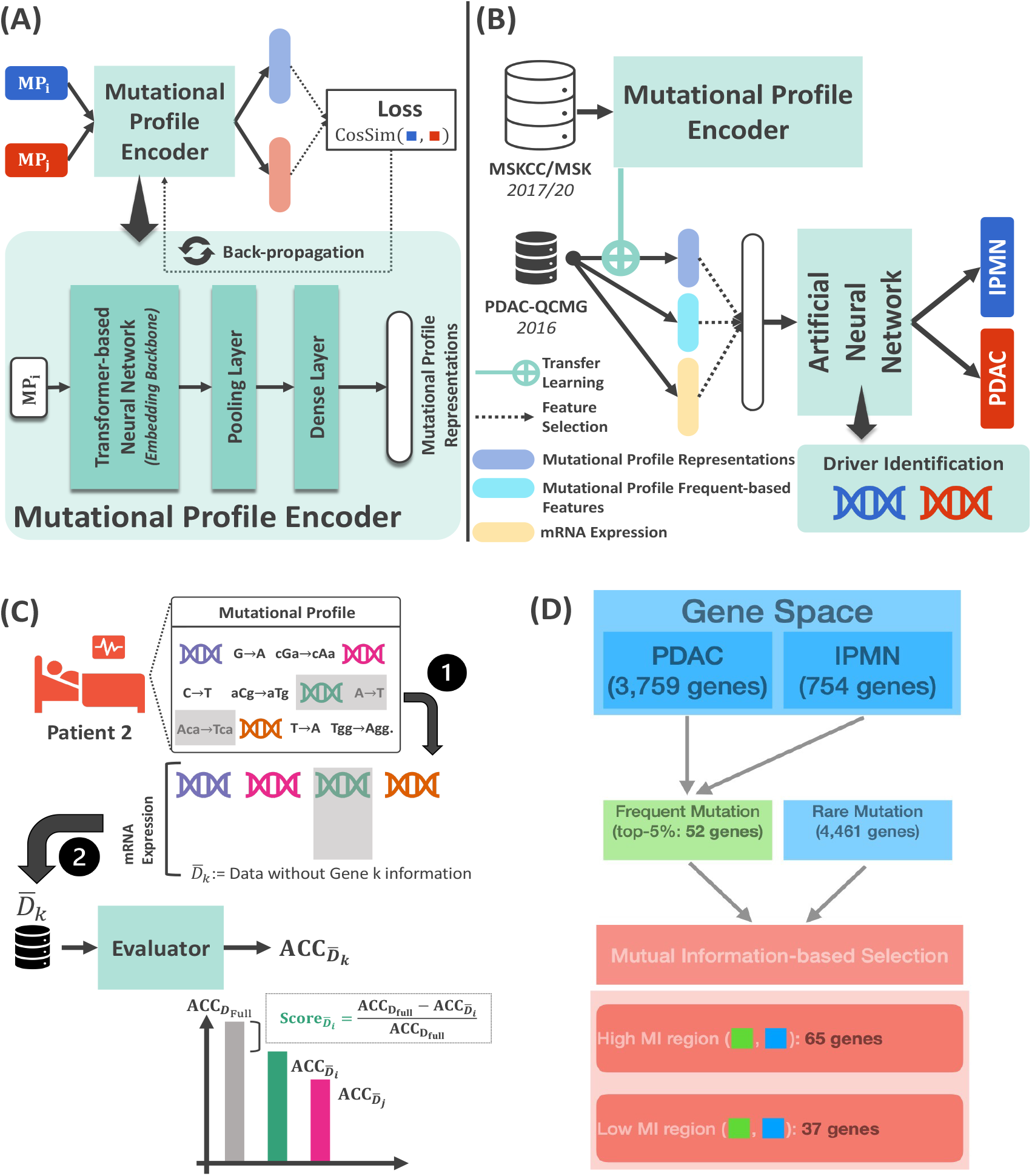
Outline of TI-MutaNET. **(A)** The mutational profile (MP) encoder is pre-trained on a large, disjointed data to obtain hidden mutational representations. The mutational profile encoder is a DL neural architecture including a Transformer-based (self-attention) backbone, followed by pooling and dense layers that decides the output feature dimensions. **(B)** The neural architecture of TI-MutaNET includes a mutational profile encoder and an adaptive-residual ANN for classification/identification tasks. The mutational profile encoder is used to extract mutational features that are incorporated with (1) frequency-based features derived from bag-of-word methods and (2) mRNA expressions for ANN classifier. The objective of TI-MutaNET is stratification between IPMN and PDAC and identification of their genetic biomarkers. **(C)** Masking mechanism and AI-driven biomarker scoring method. We randomly masks information of gene including mutational profiles and mRNA expression to create evaluated dataset. **(D)** Summary for pre-selection of scored biomarker candidates.

The second step is building a DL diagnosis system for targeted tasks using the pre-trained mutational profile encoder and an ML classifier, here adaptive Artificial Neural Networks (ANN) (**Figure 2(B)**). In general, numerical features extracted from the encoders can be analyzed by any available ML model, not restricted to ANN. However, due to its predictive potential, we demonstrate the effectiveness of mutational signatures extracted by the proposed framework on ANN. We use the adaptive ANN with residual connections, allowing deeper models with easier optimization; thus, larger model capacity with a higher predictive ability and better trainability. The pre-trained mutational profile encoders extract numerical mutational signatures from the text-based mutational profiles in a targeted cohort, independently of the pre-training sets in the first stage.

Additionally, we leverage a frequency-based feature extraction method, bag-of-word, to extract frequent statistics from the text. The final ANN classifier is trained under supervised learning to classify IPMNs and PDAC and identify subtypes of IPMNs and biomarkers for IPMN’s progression to PDAC. Since the mutational fingerprints extracted by the proposed mutational profile encoders provide numerical-mutational signatures, we combine them with other available expression datasets (**Figure 2(B)**) to increase the models’ accuracy.

#### b) Datasets

We demonstrate the application of the TI-MutaNET model to stratification of IPMNs vs. PDAC on the level of frequently and rarely mutated genes using three databases: the MSK-IMPACT targeted sequencing cohort MSKCC-2017(25), MSK-2020(26), and the whole genome sequencing data from the Queensland Centre of Medical Genomics QCMG-2016(27). All databases are annotated with binary labels, including IPMN (target = 0) and PDAC (target = 1), curated from the corresponding clinical studies. The MSKCC-2017 and MSK-2020 are used for the pre-training task (**Figure 1** (**C)**), while the QCMG-2016 set is used for the targeted task (**Figure 2(B)**). We deliberately use databases without overlapping samples to emphasize the generalization of the framework. We use a targeted cohort of patients from QCMG-2016 with fully reported mutational profiles and mRNA expression for the classification and biomarker identification task. Such data content is common in the biomedical field yet challenging for modern ML approaches due to the natural imbalance in class distribution. Here PDAC:IPMNs = 8:1. Links to the databases are provided under **Data Availability**.

The predictive robustness of the derived here gene panel (**Table 1**) was validated using additional six paired normal and tumor IPMN samples from Moffitt Cancer Center (MCC). These tumor and adjacent normal pancreatic tissue samples were harvested from patients with IPMNs between May 2017 and March 2020. These patients, diagnosed with resectable pancreatic lesions between May 2017 and March 2019 at MCC, were recruited to participate in two complementary Institutional Review Board approved studies known as the Total Cancer Care Protocol and the Florida Pancreas Collaborative study (32, 33). The MCC set includes six tumor and normal paired IPMN samples. The advantages of paired samples are to provide the opportunity to disentangle the individuals’ heterogeneity, i.e., disease intrinsic patterns can be considered.

**Table 1.**
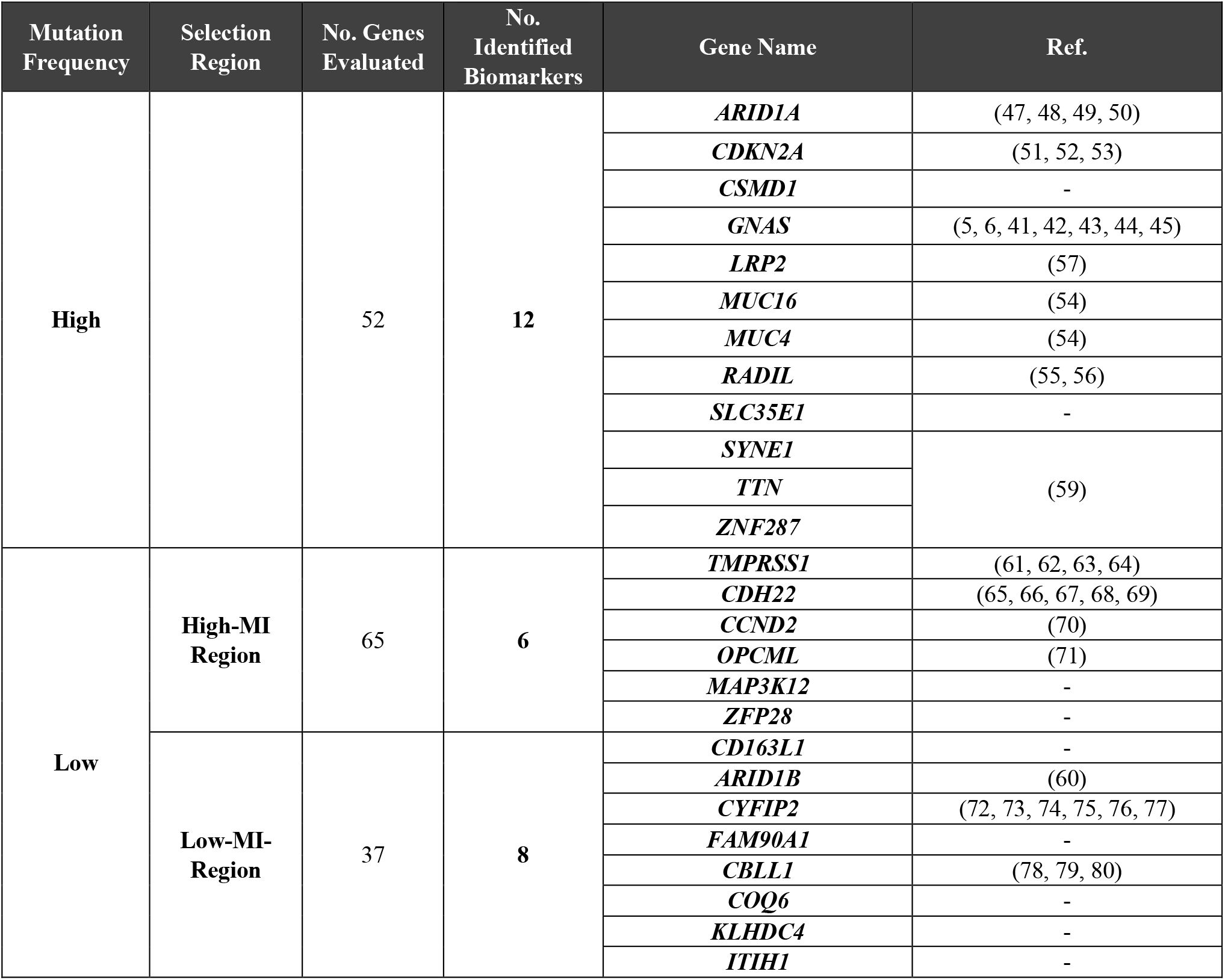
Biomarkers stratifying PDAC vs. IPMNs from frequently mutated and rarely mutated genes. Mutual Information *(38)* (MI) is applied to filter potential candidates from rarely mutated genes and reduce the initial set of 4,461 to 65 in high- and 37 in low-MI regions.

### 2.2) Framework Details

#### a) Generic Transformer Neural Architecture

The neural architecture of the generic transformer is given in (34); however, we introduce the model under the context of analyzing mutational profile sequences. Given a mutational profile of n tokens (or words) (T_1_ T_2_ … T_n_) the numerical embedding of this mutational profile is given by the matrix **X** ∈ ℝ^n×d^, where d is the pre-defined embedding space’s dimension. The generic transformer-based models aim to learn three special matrices: (1) Query := **W**_Q_, (2) Key := **W**_0_ and (3) Value := **W**_1_. The output numerical embedding of the analyzed mutational profile is given by:

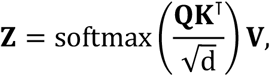

where **Q** = **XW**_Q_, **K** = **XW**_K_, **V** = **XW**_V_, and d is the corresponding dimension of queries and keys. The output representation **Z** is called single-head attentions in the transformer model. Such computations can be repeated to obtain multi-head attention, which enables capturing multiple complex contextual information from a single input.

Every element in **X** is attended in the computation of **Z** through **Q, K**, and **V**. Specifically, we compute the attention score by applying the SoftMax function on the scaled dot product 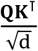, which produces probabilistic coefficients. The output **Z** is computed as the weighted sum of the values **V** = **XW**_V_, depending on every other token in the input mutational profile (**X**). Thus, the extracted vectors **Z** from the mutational encoders contain long-range dependency between gene-, nucleotide-, codon-mutations and all other dependencies (such as gene-gene, nucleotide-nucleotide, codon-codon, and all different permutations of tokens).

#### b) Self-Attention Mechanism

The self-attention mechanism is another way to illustrate how transformer-based models process input data and enable every token to attend to every other token within the same input sequence(35). Specifically, given a mutational profile sequence of n tokens, self-attention neural networks consider a complete graph 𝒢 = (𝒱, ℰ) in which 𝒱 is the nodes’ set of tokens and ℰ is the edge set connecting nodes. In other words, token T_(_ in the mutational profile sequence (T_1_ T_2_ … T_n_) is embedded into the node v_i_ with associated node feature **x**_i_. A complete graph is a dense graph where every node (or vertices) relates to another node. The graph structure is encapsulated within adjacency matrix **A**, which equals to **A** = **11**^T^. Hence, the set of neighboring nodes for any node v is the entire graph, say 𝒩_v_ = 𝒱. The learned features **z** (or representations) of node v are derived by the self-attention operator (35), given by

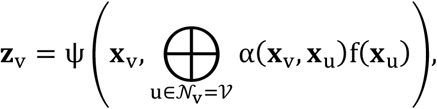

where f(.) and Ψ(.) are neural transformations (simple ANNs), and α(**x**_v_, **x**_u_) ∈ [0,1] is the attentional (probabilistic) coefficient between node u and v. The attentional coefficient indicates how much relative importance between nodes and describes the interactions among tokens. By designing a complete graph, we can capture the globally contextual information from mutational profile sequences, revealing useful patterns for prognosis. This implication is consistent with contextual embedding’s computation of **Z** given in the previous section.

#### c) Positional Encoding and Ordered Mutational Profiles

The transformer-based models or self-attention mechanisms cannot model the order of words in mutational profile sequences; the explanation for self-attention via a complete graph shows that the learning mechanism captures no convolution or recurrence, i.e., the order of nodes is not quantified in the model. Thus, in practice, positional encoding is an additional step to embed such missing positional information into the input sequences. The common choice for positional encoding is based on sine, or cosine functions with different frequencies (34). We propose periodically structuring mutational information in this framework to form mutational profile sequences. Specifically, mutated genes are in 3k, nucleotides in 3k + 1, and amino acid mutations in 3k + 2 (k = 0,1,2, ….) location of the sequence. This embedding scheme allows us to implicitly inject positional information of gene-, nucleotide-, and codon mutations by distributing their information periodically throughout the input mutational profiles. Given an input mutational profiles 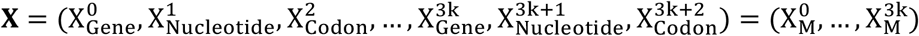 where 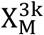 presents the mutational event 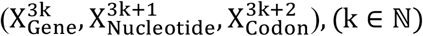.

#### d) Robust Biomarker Identification

Biomarkers identified by AI neural networks obtained by training on many samples are data-driven; within, cancer markers are attributed to influential features. Of note, the features can be interpreted by a post-hoc explanation (36), which considers the feature’s effects (masked features) on the performance of trained models. We define that the feature’s importance positively correlates with the accuracy gained by DL models. We generate data with excluded Gene k information by masking the corresponding mutational information 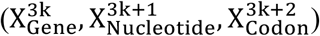 for all samples in the dataset. Besides, gene expression data for the gene are excluded for further analysis (**Figure 2(C)**); i.e., we exclude the information from Gene k to measure its importance (produced 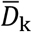 dataset), assuming it positively correlates with the accuracy gain when knowing the gene’s information *D*_full_. The final score is given by:

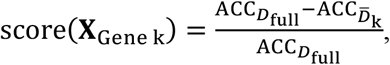

where *D*_full_ and 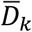 are experiments with and without the information of gene *k*. This quantity indicates how informative gene *k* is to model predictions for disease stratification or other tasks.

The proposed ANNs used in TI-MutaNET have adaptive model complexities, where their width and depth can be tuned as hyper-parameters to yield optimal configurations. By evaluating the best model setting for each quantified 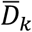 and comparing their accuracy gains, the biomarkers identified here are more robust than most post-hoc model explanation methods (37). In other words, conventional approaches perform post-hoc analyses on a well-trained model on the entire dataset. Thus, these approaches do not guarantee optimal neural solutions for each shuffle, perturbed or masked dataset. In contrast, our framework compares optimal model architectures for each scored candidate marker; thus, more robust, and reliable inferences can be drawn.

#### e) Strategy to Pre-Select Candidate Markers

##### Frequently mutated genes

We score hyper-mutated genes based on mutation profiles stratified by classes. We compute the mutations based on all different subtypes specified by genes. The number of mutated genes in mutational profiles of PDAC is 3759, and in IPMNs is 754. We considered genes with six or more mutations in PDAC and two or more in IPMNs. Disease stratification and selection of frequently mutated genes mitigate the biases from imbalanced datasets. With the threshold of mutation counts, approximately 5% of mutated genes (**Figure 2(D))** from each disease are chosen for evaluation. As a result, we select and evaluate 52 genes (with fully reported gene expression data), equivalent to 52 datasets with the information about Gene masked.

##### Rarely mutated genes

Our proposed biomarker scoring method requires global optimization of AI models, including model architecture and weights (hard-coded knowledge). Thus, scoring all rare mutations demands massive computational resources (data abundance in the rarely mutated spectra is a challenge, as 4,461 genes were extracted from the QCMG-2016 database). We reduce this cost by post-selecting potential candidates by mutual information (MI) (38) shared with the already scored markers. This means we can evaluate two distinctive regions: 1) genes sharing high MI (relevancy) and 2) genes sharing low MI with scored genes (redundancy) (**Figure 2(D)**). This approach is our adaptation of maximum relevancy and minimum redundancy to eliminate less informative regions (39). With the strategy proposed, we reduce 4,461 genes (mutations in PDAC or mutation in IPMNs, remaining 95% of mutated genes) to 102 genes for the actual gene scoring stage.

#### f) Evaluation protocol

To evaluate our model performance, we avoid using the conventional accuracy metric defined by

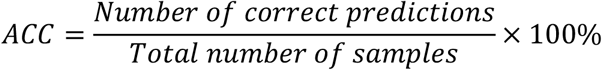

since the result could lead us to misleading comparisons and further evaluations. Specifically, our investigating data scenario has a proportion of positive class (PDAC) of about 89%. Thus, random classifiers can reach nearly 90% with non-robust inference. We use the area under the curve (AUC) to address this issue since the metric measures the model’s ability to discriminate between positive (PDAC) and negative (IPMN) classes with a range of thresholds. In our case, the metric overcome the biases from imbalanced data distribution by incorporating the true positive rate (TPR) and false positive rate (FPR) by different thresholding values. Mathematically speaking, the computation for AUC is given by:

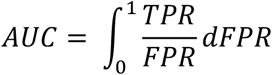

where:

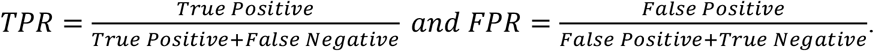

## 3) Results

### 3.1) TI-MutaNET enables Highly Performed ANN Classifiers for Stratification between IPMNs vs. PDAC

We calculate the normalized biomarker scores (BS, between [0,1]) for 52 frequently mutated genes from TI-MutaNET. Each mutational profile encoder produces different BS landscapes with varying marker scores from ALBERT, BERT, and MiniLM-L6, while BS from MiniLM-L12 is nearly uniform. We enhance the inference robustness by averaging BS from the four variants of the proposed models. We use two statistical quantities, the p-value of the t-test between diseases’ mRNA expressions and their variance for post-selection of cancer markers (**Figure 3(C)**). P-value reveals the statistical significance of such gene expression from a targeted cohort to stratify diseases, while the remaining criteria present feature variations across all patients. However, such a statistical test could make a non-robust inference in the low-limit data set with imbalanced data distribution. Specifically, *MUC4, SYNE1*, and *ZNF287* can be addressed as biomarkers under the t-test with a statistical significance under *α* = 0.1, while *TTN* has a statistical significance of *α* = 0.05 in the stratification of IPMN vs. PDAC. However, such a univariate test fails to address commonly known biomarkers of PDAC, such as ARID1A (p-value = 0.19), CDKN2A (p-value = 0.74), and GNAS (p-value = 0.3). In contrast, our biomarker score depends on multivariate analysis, including multiple modes such as base pair mutation, codon mutation, gene mutation, and mRNA expression. Thus, our research could lead to more robust and reliable biomarkers. We only consider genes with BS greater or equal to 0.5 for the final biomarker set.

**Figure 3:**
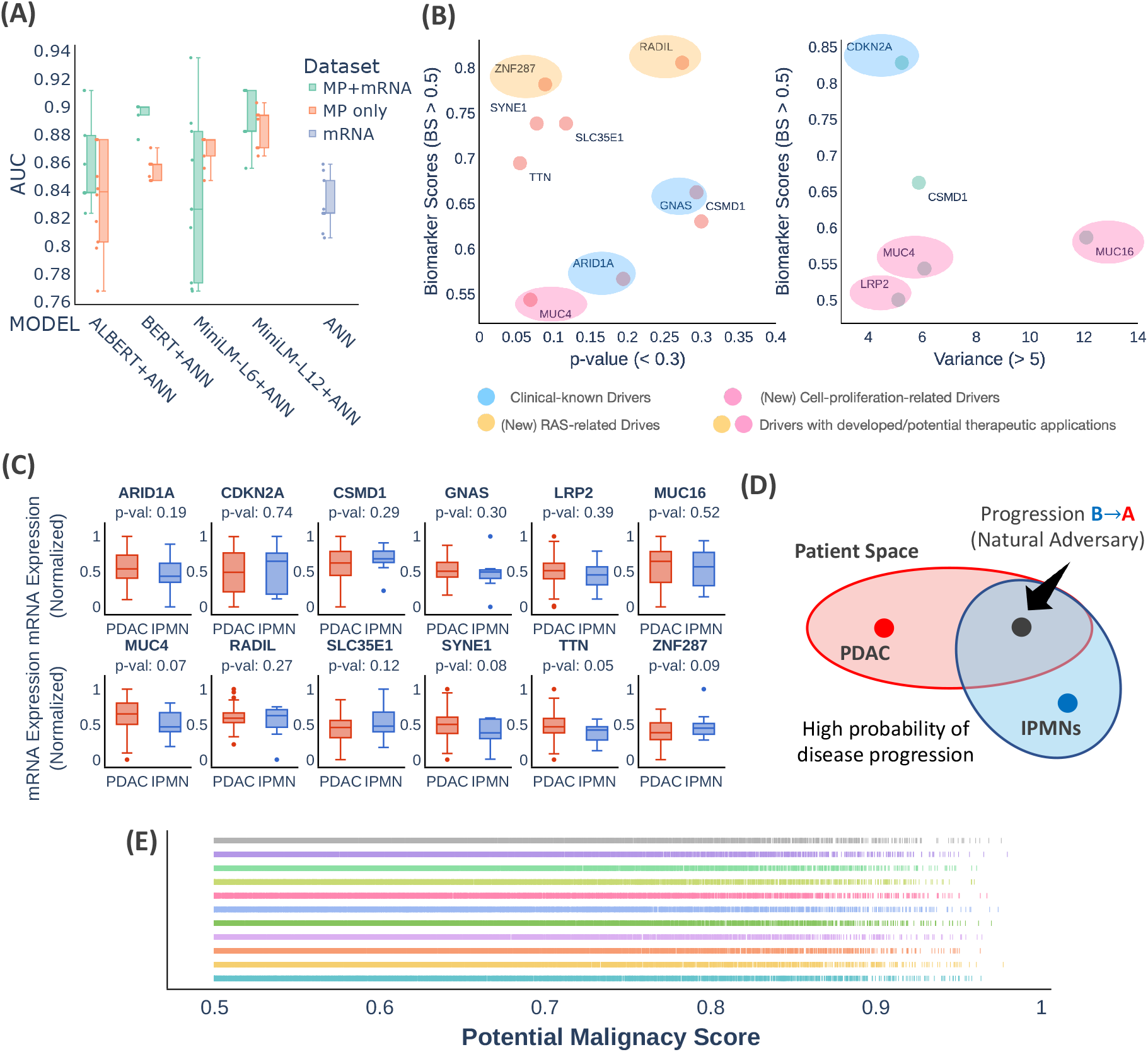
Classification and biomarker identification results of TI-MutaNET. **(A)** Classification results of TI-MutaNET based on top-10 AUC scores out-of-100 random stratified evaluations. All databases are annotated with binary labels, including IPMNs (target = 0) and PDAC (target = 1), curated from the corresponding clinical studies. Identified genetic biomarkers for stratification of IPMNs vs. PDAC based on the post-selection of statistical significance (p-value) and variance of mRNA expression. **(C)** The MP-derived gene panel from this study - includes 12 genes with their statistical significance of mRNA expressions and was curated using the QCMG-2016 dataset. Only *MUC4, SYNE1, TTN*, and *ZNF287* are detected as gene-expression-derived biomarkers by t-test with statistical significance under *α* = 0.1. **(D)** Natural adversary analysis for progression pathway from IPMNs to PDAC - based on the hypothesis that IPMN tumors with a high probability of progression will share similar genetic patterns with PDAC. **(E)** The malignancy score - based on an ensemble of models with optimized model architecture and weights.

Classification results of TI-MutaNET, based on the top 10 AUC scores of 100 random stratification evaluations with four mutational profile encoder variants, are shown in **Figure 3(A)**. Features extracted from the transformer-based models helped reach an average of 0.86 in AUC with the ALBERT model and a maximum of 0.88 in average AUC with MiniLM-L12, compared to 0.82 in average AUC using mRNA data only. These gains are significant regarding robust accuracy metric (AUC) for problems with natural imbalance in the data distributions such as PDAC and IPMNs (8÷1 in QCMG-2016). When combined, the mutational features and corresponding mRNA expression data derive higher predictive ML classifiers with an accuracy of approximately 0.94 in AUC with MiniLM-L6+ANN. These numerical results indicate that mutational fingerprint vectors from the proposed mutational profile encoders represent quantified diseases, enabling more powerful DL classifiers. The generalization of our framework is shown by using different databases for pre-training the mutation encoders and training for the main tasks. This indicates that DL approaches allow the development of highly accurate models for a targeted cohort with a small number of samples by leveraging knowledge from externally related databases.

The representative features of proposed MP encoders induce more accurate DL classifiers – above 0.82 of AUC in all variations of mutational profile encoders. Moreover, consistency in the performance improvement across all transformer-based encoders supports the concept of treating mutational profiles as a language of the human genome, in which the information can be efficiently retrieved and analyzed using a self-attention mechanism (**Figure 3(A)**). Thus, we see the great potential of neural architecture applications in quantifying the mutational signals in cancer research. We believe the proposed framework is an initial attempt to deconvolute mutational signals with transformer-based language models.

Large models do not guarantee optimal neural solutions for mutational profile encoders. The extremely complex BERT models with 110 million parameters lead to a degradation of accuracy compared to significantly smaller models such as MiniLM-L6 and MiniLM-L12, with only 22.3 and 33 million parameters, respectively. This phenomenon is commonly seen when training highly complex models with small datasets that may result in local optimal of the neural solutions. However, we claim an optimal neural architecture exists for mutational profile encoders depending on specific input datasets (40). The selection and design of the optimal mutational profile encoders for data-specific problems are worth investigating in future studies.

### 3.2) TI-MutaNET Suggests Clinically Relevant Biomarkers for Stratification of IPMNs vs. PDAC

#### Frequently Mutated Genes

Twelve data-driven biomarkers for stratification of IPMNs and PDAC are discovered by TI-MutaNET (**Figure 3(B)** and **(C)**). Several identified genes, including *GNAS* (BS = 0.63), *ARID1A* (BS = 0.57), and *CDKN2A* (BS = 0.83) are consistent with the clinically known regime; GNAS mutations commonly occur in the majority of IPMNs (5, 6, 41, 42, 43, 44, 45). These guanine nucleotide-binding proteins (G-proteins) are transducers for many signalling pathways controlled by the G protein-coupled receptors (GPCRs) (46). Interestingly, *GPCR98* is identified as a highly significant biomarker (BS = 1) by two variants of TI-MutaNET, including MiniLM-L6 and MiniLM-L12.

*ARID1A*, involved in the transcriptional activation and repression of selected genes by DNA-nucleosome topologic alterations, is reported as a frequently mutated gene in PDAC and IPMNs (47, 48, 49). The simultaneous loss of this biomarker and *PTEN* in mouse pancreatic ductal cells leads to intraductal tubulopapillary neoplasms, which can progress to PDAC (50). *CDKN2A*, clinically identified as a common PDAC biomarker (51, 52, 53), is a negative regulator of normal cell proliferation. This gene strongly interacts with *CDK4* and *CDK6*, the binding partners of cyclones in the hyper-phosphorylation of retinoblastoma within the cell cycle.

Several biomarkers identified by TI-MutaNET are associated with clinically known biomarkers in IPMNs, including *MUC4* (BS = 0.54) and MUC16 (BS = 0.59). These genes could promote tumor growth and progression by repressing apoptosis instead of cell proliferation (46). A case study on a resected two-branch duct-type IPMNs showed concurrent activation of genes in the same family (*MUC5AC* and *MUC6*), for high-grade dysplasia(54). Thus, we believe that an extension to study the whole *MUC* family could reveal biological evidence for IPMNs grading, which is worth exploring in future research.

The remaining identified biomarkers seem novel or rarely known. *RADIL*, a *RAS*-associated and dilute domain-containing protein, is a potential marker for IPMNs vs. PDAC stratification (BS = 0.8). This gene interacts with *RAS*-related proteins, such as *RAP1A, RRAS2, RASGEF1C*, and *HRAS*. It is noted that *HRAS* is an alternatively mutated *RAS* isoform (46) and can be considered an intrinsic biomarker for PDAC. Besides, *RADIL* is identified as a putative activator and a binding partner of other *RAS* isoforms indicating multiple interactions with *KRAS* (46). *RADIL* also plays a vital role in the cell adhesion process, regulated by G-protein beta-gamma subunits (55) that are potential targets for therapeutics (56).

The TI-MutaNET proposes *CSMD1* (BS = 0.66), a potential suppressor of squamous cell carcinomas, as a marker for the stratification of IPMNs vs. PDAC. *CSMD1* could interact with *SLC35F2*, a putative transporter in the solute carrier family 35, with discovered *SLC35E1*(46) (BS = 0.74). The *LRP2* is another identified marker crucial in producing receptor protein megalin. It is worth noting that this gene was previously recognized in prostate cancer and disorders in lipoproteins and cholesterol in the blood (46). Polymorphisms of *LRP2* can increase the activity of megalin, leading to more aggressive tumor growth (57). The biomarkers with substantially higher BS scores and p-value under 0.1 are *TTN* (BS = 0.7), *SYNE1* (BS = 0.74), and *ZNF287* (BS = 0.78). *TTN* is a potential partner with E3 ubiquitin-protein ligase *TRIM63*, which could interact with *SMAD3* and *SMAD4* (46), recognized *PDAC* biomarkers. The marker also plays a role in maintaining muscle mass (58), a significant factor in cancer cachexia. *ZNF287* could potentially interact with *RASAL2* – the regulator of *RAS*-cyclic *AMP*, based on the literature (46). Importantly, *RASAL2* is involved in the interactions with multiple *RAS* isoforms, including *KRAS, HRAS, NRAS*, and *NF1*, an associated biomarker in many cancer types (59). We summarize all identified markers with their clinical significance in cancer, PDAC, and IPMNs in **Table 1**.

#### Rarely mutated genes

We broaden the scope of our analysis to rarely mutated genes, which are commonly ignored due to the weak signals in statistic counts. Since the transformer-based models learn local contextual information (e.g., only codon mutations) and global contextual information from the input sequences (e.g., relative information between each information token or word), thus, our approach can more efficiently identify biomarkers from rare mutational signals. Under the rarely mutated genes, we discover six and eight potential biomarkers for stratification of PDAC vs. IPMNs from high- and low-MI selection regions, respectively (**Table 1)**, with several interesting genetic alteration patterns observed. First, the biomarker *ARID1A* is found within the highly mutated areas; however, its mutually exclusive paralog, *ARID1B*, located in rarely mutated regimes with less relevance to already scored genes, is also identified as a biomarker. We find eight patients with *ARID1A* mutations: seven samples are PDAC, and only a single case is IPMN. The mutation from *ARID1B* is recorded in mutually exclusive selection. Our model reveals potential markers that could be omitted by analyses using conventional ML models due to the weak input signals. The concurrent loss of *ARID1A*/*B* augments cancer development and disruptive redistribution of BAF complex–chromatin reconfiguration of *SWI/SNF* mutated in 20% of cancer types (60).

Several notable biomarkers from our analysis, although ignorable by statistical counts due to rare mutations, have significant clinical evidence for potential markers of carcinogenesis and malignancy. For example, *TMPRSS1* (or Hepsin) and its type II transmembrane serine proteases *TMPRSS3*, are associated with malignancies such as, e.g., *breast* cancer(61); Moreover, *TMPRSS9* affects PDAC cells by pro-uPA activation (62) while *TMPRSS4* stimulates cell proliferation in PDAC and inhibits apoptosis by activating *ERK1/2* pathways (63). This is an important genetic family in cancer research. Type II transmembrane serine proteases (TTSPs) are markers in several tumor types (64). *CDH22* is a protein-coding gene related to ERK signaling, which regulates cell behavior and fate (65). This biomarker is also used with *RADIL* and *SLC35E* as the targeted genes of long non-coding RNAs *MIR600HG*, serving as a potential marker of *PDAC* (66). Besides, the marker has been used for identifying malignant progression of epithelial tumors such as colorectal (67), breast (68), or melanoma (69) cancers suggesting its potential as a PDAC progression marker. Based on our study’s statistical evidence, *CDH22* could be a potential marker for the progression pathway from IPMNs to pancreas malignancy. The cyclin D2 (*CCND2*), which encodes for proteins that regulate *CDK* kinases during the cell cycle (46), can also play a significant role in the stratification of malignancy (PDAC) vs. pre-malignancies (IPMNs or other cysts). It was found that *CCND2* mutations occurred in 0.63% of gastric carcinoma patients (70). Another study of circular tumor DNA-based tests on 282 patients with PDAC at Mayo Clinic reveals that mutations of *CCND2* and common markers *KRAS, SMAD*, and *TP53* are frequent in the cohort with advanced disease (81). The cyclins in the same family, *CCND1* and *CCND3* (or *D1* and *D3*), are commonly overexpressed at the same time (co-overexpressed) in PDAC (82). Suppressing *CCND3* reduces the phosphorylation of retinoblastoma protein or cyclin A produced in the cell cycle. Besides, *CCND1* interacts with *MAPK* and *NF*-B signaling, suggesting that this biomarker suppressed *PANC1* cells. This biomarker also can detect IPMNs with PDAC, as indicated in a real-time targeted genome profile analysis (83). A combination of *CCND2* with *FOXE1, NPTX2, PENK*, and *TFPI2* is considered highly predictive of pancreatic tumors, with nine over eleven patients with tumors but without pancreatic neoplasia. The signal on *CCND2* biomarker, also used in the ctDNA test on 104 patients (84), suggests two things. First, mutations of *CCND2* take third place just after *KRAS* and *TP53* in the quantified cohort among the 39 patients with stage III (local-advanced PDAC) and 65 with stage IV (metastatic disease). Second, such mutations of *CCND2* significantly reduce the median progression-free survival to 3.7 months, compared to 8.2 months in non-mutated samples. We also found a tumor suppressor *OPCML*, which is already recognized in epithelial ovarian cancer (71, 85). Moreover, a new potential inhibitor, *CYFIP2*, which promotes cell growth, cell migration (72), and chemoresistance in pancreatic neuroendocrine neoplasms cancer (77) by activation of the Akt signal (73), showed potential for therapeutic applications among other malignancies (74, 75) or metastasis cases (76).

### 3.3) Estimating Malignancy Score with Philosophical Concept – Natural Adversary

We propose a new application of the natural adversary concept to analyze the progression from IPMNs to PDAC in **Figure 3(D)**. We define adversarial-biological samples to ML models as observations that are challenging to be classified by the models. In our hypothesis, IPMN samples will have a high probability of disease progression due to similar genetic (hidden) patterns to malignant samples. We reused trained AI models obtained during optimization for neural architectures. With four backbone NLP neural architectures and 11,750 models trained with a random stratification strategy, we post-select effective models only, i.e., excluding the models with nearly random predictions (7,850 models). By the neural architecture design, the ANNs will output the probability of being a PDAC sample (by applying SoftMax) ranging from 0 to 1 (AI-driven malignancy score from [0.5,1] is shown in **Figure 3(E)**. Random stratification evaluation is a synthesized method to provide different train/test data distributions for models, enabling a more generalized inference than a stand-alone ML classifier.

Natural-adversarial samples are originally addressed in computer vision applications, referring to input images that can deceive ML models (86). For example, our previous work (40) addressed melanoma lesions are the natural-adversarial example of skin lesions, in which the deadliest skin cancer is commonly share similar visual features to benign lesions. Similarly, in the context of genetic data in IPMN vs. PDAC, adversarial examples will have a great chance of making AI models providing false predictions. Hence, the prediction confidence on these samples will be low, i.e., inclining probability toward the malignancy class (in binary case only). Therefore, we use these patterns to detect IPMNs examples with a high risk of progression to PDAC. The proposed conceptual model is a cost-efficient method to recycle many expensively trained models to understand disease mechanisms better. Based on the above natural adversary analysis, five of the 11 IPMN samples in the QCMG-2016 database show a high likelihood of progression. These include patients 1, 2, 6, 7, and 10 that have the score diffused in the region [0.9-1] (**Figure 3(E)**), which suggests a higher chance for progression.

### 3.4) External Validation of the AI-derived Biomarkers

Only one biomarker – *TMPRSS1D* (**Table 1**) – is excluded from this analysis (not recorded in the MCC dataset). We compare the predictive power of the AI-derived markers with clinical panels in (6, 8) on MCC data under a leave-one-out evaluation: we train a logistic regression on five sample pairs and test on the remaining pairs of a single individual. **Figure 4(A)** shows that our panel has comparable accuracy to the two clinical panels. Specifically, we use three accuracy metrics, including Accuracy (ACC), Area under Curve (AUC), and F1-score (combination of precision and recall), for evaluation.

**Figure 4.**
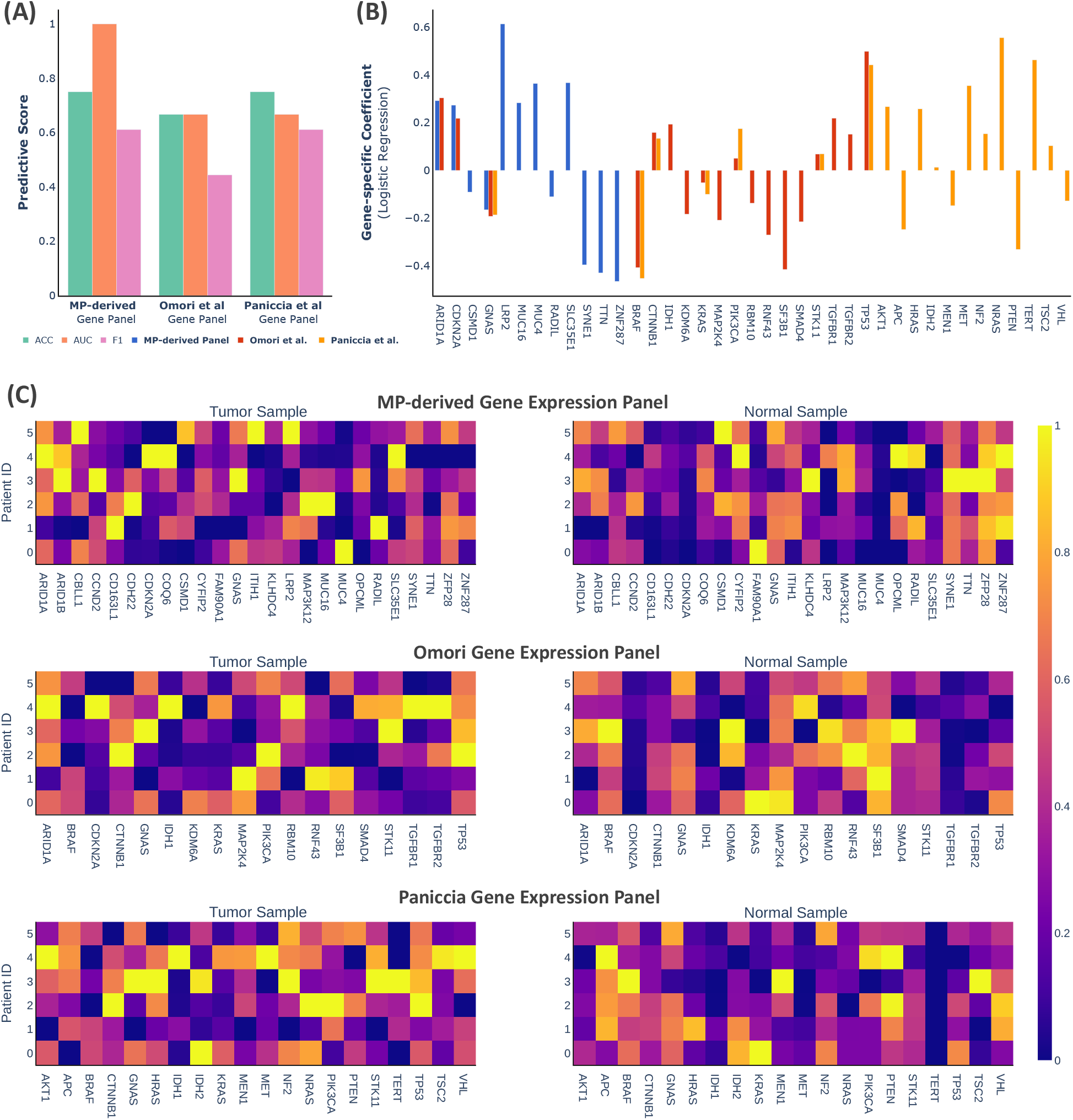
External validation of the MP-derived gene panel on MCC data, including six pairs of tumor-normal IPMNs. (**A**) Comparison of the gene panel developed here with the clinically derived panels (6, 8). (**B**) Coefficient logistic regression analysis under leave-one-out evaluation using MCC data. (**C**) Heatmaps of normalized RNA expressions across three panels, taking value from [0,1].

Our predictive, mutation-derived gene panel has a higher AUC (0.82) compared to Omori’s (6) or Paniccia’s (8) panel (0.66 in both panels). In contrast, Paniccia’s panel achieves the highest ACC of 0.75 (surpassing 0.66 from our panel and Omori’s). Paniccia is also characterized by a higher F1 score than the other two panels. This means that our attempt to leverage optimal data collection from publicly available datasets to analyze the MCC dataset better led us to an equivalent genetic panel for stratification of pre-malignancy/malignancy for IPMNs samples without any cost for clinical protocol. Besides, we discover several potential biomarkers not addressed in clinical literature. Specifically, the two panels from (6, 8) share eight overlapped genes, while our framework produces only three overlapped markers (**Figure 4(B)**).

**Figure 4(C)** shows several interesting patterns of tumor-normal samples: *CYFIP2, SYNE1, TTN, ZFP28*, and *ZNF287* are under-expressed in tumor samples [0.4-0.6], while over-expressed in normal tissue [0.8-1]. Similarly, we find that *SF3B1, TP53*, and *MAP2K4* have the same contrariness in expression, in which expressions are low in malignant samples and higher in pre-malignant samples. Distinguishable patterns were also found in Paniccia’s panel, including *AKT1, APC, BRAF, CTNNB1, NF2*, and *NRAS*. Based on these findings, we propose that our AI-driven gene panel, being as effective, is more cost-efficient compared to the other panels since the cost for such a diagnostic panel is the computational cost of trainng AI models on public avaiable databases (without clinical trials).

## 4) Conclusions

In summary, we introduce the TI-MutaNET model, an AI-enabled cancer stratification and biomarker identification framework. We demonstrate the proof-of-concept for the proposed neural pipeline using pre-cancerous cysts (IPMNs) and PDAC. We address the insufficiency of current approaches by integrating information from mutational profiles and propose model learning mechanisms that help formulate mutational sentences by periodical positional encoding. We provide an in-depth illustration of the model through its architecture and data-centric perspectives. The proposed data configuration allows transformer-based models to address human mutational profiles efficiently and implicitly, casting positional relationships over the inputs. Our study results in the identification of the new biomarkers stratifying IPMNs vs. PDAC. We also introduce a practical method for maximizing AI models’ utility to understand disease progression better. We find that recycling of the AI models can further support cancer research, and we hope the proposed technique could motivate further studies on optimizing the current AI approaches. Furthermore, although the application of TI-MutaNET is demonstrated on pancreatic malignancies, we believe the model can be generalized to study other diseases and their genetic causes.

## Data Availability

The datasets were obtained via https://www.cbioportal.org/:
https://www.cbioportal.org/study/summary?id=msk_impact_2017,
https://www.cbioportal.org/study/summary?id=msk_ch_2020,
https://www.cbioportal.org/study/summary?id=paad_qcmg_uq_2016.

https://github.com/namnguyen0510/TI-MutaNET

## Nomenclature

Abbreviation: Meaning
TI-MutaNET: Text-based Integration of Mutational Profiles for Classification and Biomarker Identification
AI: Artificial Intelligence
ML: Machine Learning
DL: Deep Learning
IPMNs: Intraductal Papillary Mucinous Neoplasms
PDAC: Pancreatic Ductal Adenocarcinoma
ANN: Artificial Neural Network
QCMG: Queensland Centre of Medical Genomics
MSKCC: Memorial Sloan Kettering Cancer Center
MCC: Moffitt Cancer Center
CL: Contrastive Learning
RL: Representation Learning
TL: Transfer Learning
AUC: Area Under Curve

## Data Availability

The datasets were obtained via https://www.cbioportal.org/.

Link for each dataset (pre-processing):

- MSKCC-2017: https://www.cbioportal.org/study/summary?id=msk_impact_2017,
- MSK-2020: https://www.cbioportal.org/study/summary?id=msk_ch_2020,
- QCMG-2016: https://www.cbioportal.org/study/summary?id=paad_qcmg_uq_2016.

## Code Availability

This work’s source code is available on GitHub at: https://github.com/namnguyen0510/TI-MutaNET.

## Notes

### Competing Interest Statement

The authors have declared no competing interest.

### Funding Statement

This study did not receive any funding

### Author Declarations

https://www.cbioportal.org/study/summary?id=msk_impact_2017 https://www.cbioportal.org/study/summary?id=msk_ch_2020 https://www.cbioportal.org/study/summary?id=paad_qcmg_uq_2016

